# Facile Filtration-Based Workflow for Facilitating Sensitive Detection of Pathogenic Bacteria from Blood

**DOI:** 10.1101/2023.11.17.23298597

**Authors:** Dong Jin Park, Liben Chen, Tianqi Wu, Pei-Wei Lee, Kuangwen Hsieh, Tza-Huei Wang

## Abstract

Bloodstream infections (BSIs) are a global health concern, contributing to high mortality rates and increased healthcare costs. Current diagnostic methods, such as blood culture, matrix-assisted laser desorption/ionization time-of-flight mass spectrometry (MALDI-TOF MS), immunoassays, and nucleic acid amplification tests (NAATs), have lengthy workflow bottlenecked by the need of culture based method This study introduces an innovative blood processing method that overcomes this limitation, enabling rapid and culture-free isolation of pathogenic bacteria from whole blood. The method combines osmolysis, blood separation membrane filtration, detergent and enzymatic lysis, and bacterial capture, requiring minimal specialized equipment. Notably, it uses a small blood sample volume (0.5 mL or below), making it suitable for pediatric patients. The workflow involves four simple steps and can be completed in approximately 30 minutes, providing rapid pathogen isolation. The protocol was successfully tested with two most common causative bacteria, S. aureus and E. coli, achieving sensitive detection down to approximately 10 CFU using benchtop PCR. The culture-free approach accelerates the diagnosis process and minimizes the risk of bacterial population alterations during culturing. While further optimization and testing with a broader range of pathogens are needed, this method holds promise in advancing the diagnosis of life-threatening bacterial infections. Future applications may include rapid point-of-care testing in resource-limited settings.

## Introduction

Bloodstream infections (BSIs) are a serious global health concern, responsible for a substantial number of hospital admissions and associated with high mortality rates. The impact of BSI on patient outcomes is evident through global studies, revealing an alarming estimate of 48.9 million sepsis cases worldwide. A staggering 19.7% of all global deaths are attributed to this condition, supporting BSIs as a leading cause of infectious disease-related fatalities in healthcare settings [1]. The repercussions extend beyond mortality, as patients with BSIs often contend with prolonged hospital stays, resulting in escalated healthcare costs and an increased risk of severe complications such as septic shock, organ failure, and secondary infections [2]. Among the diverse causes of BSIs, Bacteria significantly contribute to these life-threatening infections, with a study indicating that 92% of BSIs are caused by bacteria. Within this BSI population, E. coli and S. aureus emerge as the most prevalent strains [3]. Furthermore, 9.9% of all healthcare-associated infections arise from primary bloodstream infections, accounting for an estimated 250,000 occurrences annually [4, 5]. Considering these circumstances, there is a pressing and critical need for enhanced diagnostic methods to improve the early detection and management of bacterial bloodstream infections, thereby mitigating the associated morbidity, mortality, and healthcare burdens.

Currently, blood culture is considered the standard for diagnosing bloodstream bacterial infections but is limited to culturable bacteria and suffers from lengthy turnaround time of 48 to 72 hours [6], which can delay the initiation of targeted antimicrobial therapy. Matrix-assisted laser desorption/ionization time-of-flight mass spectrometry (MALDI-TOF MS) has demonstrated high accuracy and rapid turnaround time for bacterial identification, making it valuable in guiding targeted therapy [7]. However, this method typically necessitates culturing a sufficient amount of bacteria for detection. In addition to the lengthy drawbacks of blood culturing, this method requires access to specialized mass spectrometry equipment and is unlikely to provide direct detection of bacteria in blood samples [8]. Immunoassays, such as enzyme-linked immunosorbent assays (ELISAs) and lateral flow assays, detect bacterial antigens or antibodies in the bloodstream. These tests offer relatively rapid results and can be performed at the point of care, making them suitable for resource-constrained settings. However, the sensitivity and specificity of immunoassays may vary depending on the target antigen and the viscosity of the blood environment, leading to false-negative results [9]. Nucleic acid amplification tests (NAATs) such as PCR amplify and detect specific bacterial DNA or RNA sequences, allowing for sensitive and specific identification. PCR-based tests have shown promising results in reducing the time to diagnosis compared to traditional culture-based methods, with sequencing and qPCR-HRM standing out as a gold standard for pathogen identification with high sensitivity [9]. However, these techniques may require sophisticated laboratory infrastructure and sample processing procedures for pathogen isolation, which can be a constraint in resource-limited settings. Therefore, an innovative blood processing method holds the potential to advance NAAT-based diagnosis of bacterial bloodstream infections, enabling early intervention and improved patient outcomes.

To date, numerous methods have been proposed for blood processing and bacteria separation. The direct mechanical filtration, involving the passage of blood through membrane filters, appears as the most straightforward strategy. However, it encounters challenges when white blood cells (WBC) and, especially, red blood cells (RBC) aggregate into a “filter cake,” obstructing further passage of both blood and bacteria [10]. Another approach focuses on plasma or serum separation using the dextran sedimentation method [11]. This method separates RBC from plasma through sedimentation in a dextran-enriched solution. Although effective in reducing WBCs and RBCs, its drawback lies in the requirement of additional steps for bacteria concentration and purification due to the large blood volume involved. Additionally, the presence of bacteria interacting and adhering to blood cells and platelets, such as S. aureus [12,13], presents an additional challenge in the recovery process. Moreover, coupling with highly sensitive tests like NAATs is not directly compatible, necessitating supplementary processing methods, including target capture, purification, and concentration for reliable target detection. Centrifugation combined with cross-flow filtration utilizes sedimentation velocity-based differentiation followed by size-based cross-flow filtration within a viscous solution. This is achieved through centrifugal sedimentation with an angled filter for bacterial passage [14]. However, this technique shares similar limitations, including culture-based detection, a high final sample volume (18 mL), and unknown compatibility with NAATs. The considerable final sample volume restricts quantitative bacterial analysis. In the realm of microfluidic-based pathogen isolation methods, various techniques have been explored, such as elasto-inertial microfluidics [15], acoustophoresis [16], surface acoustic wave (SAW) [17], dielectrophoresis [18], and magnetic bead-based separation [19]. While these methods demonstrate varying levels of bacterial detection sensitivity, particularly at high bacterial concentrations, they exhibit incompatibility for sepsis diagnosis. Despite microfluidic methods allowing rapid bacteria separation, they share common limitations such as low sample throughput, limited removal efficiency of cells or debris, low bacteria recovery, and the need for a specialized laboratory environment (clean room) for device fabrication. The existing literature highlights the challenges and limitations of current blood filtration methods for bacterial isolation. Therefore, the development of an innovative blood processing method is imperative to address these challenges and fulfill the criteria of efficiency, sensitivity, and simplicity for the improved diagnosis of bacterial bloodstream infections.

A novel technique that addresses the limitations of current methods would require several key elements. First, it should enable rapid and simple sample preparation directly from whole blood with minimal handling and sufficient processing capacity to capture pathogens even in low abundance while considering minimal invasiveness(<1 mL for pediatric patients) [20]. Second, an effective workflow should separate, enrich, and concentrate pathogens or target analytes from background interferences, thereby enhancing sensitivity and specificity. Third, the technique should offer sensitive, quantitative, and accurate detection with species-level identification to differentiate pathogens from contaminants or commensals, ensuring precise diagnosis and appropriate antimicrobial treatment. Here we present a culture free filtration based bacteria isolation method utilizing two distinct filters to separate and purify bacteria from blood. The filtration-based method proposed has the potential to address the limitations described previously without the sacrifice of the speed and accuracy of sepsis diagnosis by providing a rapid (<1 hour) and efficient method for pathogen separation from septic blood samples, which can be coupled with bacteria identification and antibiotic susceptibility testing. Furthermore, this method is fit for non-centralized limited resource settings while requiring minimal technical experience.

## Methods

### Bacteria Preparation

The bacterial strains E. coli (ATCC 25922) and S. aureus (ATCC 29213) were cultured on Tryptic Soy Agar supplemented with 5% sheep blood and incubated at 37 °C with 5% CO2 for 12 hours. Following incubation, bacterial colonies were resuspended in water. The concentration of colony-forming units (CFUs) was assessed using the OD600 McFarland standard, and subsequently, the bacterial suspensions were diluted to working concentrations. To accurately determine the viable CFU, plating was performed using the volumes employed for inoculation.

### Blood Preparation

To emulate a bacteremic state, 500 μL of human blood obtained from BioIVT (K2EDTA, gender unspecified, 4℃ storage) underwent treatment with 85 μL sodium polyanethole sulfonate (SPS, 0.35% in 0.85% NaCl) for 10 minutes at room temperature. This treatment served a dual purpose, promoting bacterial stability against humoral and cellular factors in the blood environment while also acting as an anticoagulant [21]. Following this treatment, E. coli ATCC 25922 and S. aureus ATCC 29213 were inoculated into the blood sample. To facilitate the release of internalized bacteria from erythrocytes, the blood was diluted 10-fold with sterile water (4500 μL). This dilution induced osmolysis, preventing the capture of internalized bacteria in the initial separation membrane.

### Filter Preparation

A Swinnex filter holder (SX0002500, Millipore Sigma) equipped with two distinct filter membranes was employed for blood filtration. The first membrane, a gradient filter designed for cell/plasma separation (International Point of Care Inc, S/G Membrane filter, 35 micron (top) - 2.5 micron (bottom)), facilitates separation of cells and large debris from sample. The second membrane, a 0.4 μm filter (Sterlitech, PCTE, 0.4 micron, 25mm), is dedicated to capturing bacterial species. To ensure a sterile environment, both the filter holder and filter module underwent cleaning with 10% NaClO and 75% EtOH before autoclaving.

### Assay

The filtration assay began with the 5 mL osmotic-lysed blood poured into a 5 mL syringe then pushed through the cell/plasma separation membrane with gradient pores to capture remaining erythrocytes. The resulting filtrate, containing bacteria, was collected in a sterile 15 mL Falcon tube. Subsequent steps involved incubating the filtrate with 500 μL of 0.1% CHAPS (3-((3-cholamidopropyl) dimethylammonio)-1-propanesulfonate) for 4 minutes followed by a 1-minute incubation with 500 μL of 0.5% trypsin-EDTA at room temperature for residual blood cell digestion. The bacterial cells were then captured on a 0.4 μm filter membrane by flowing digested samples through capture filters using a 5 mL syringe, subsequently rinsed with 4 mL of sterile water to eliminate any residual blood or plasma components. The captured bacteria from the filter membrane was eluted with 1 mL 0.1% tween 20 in water by pushing the elution buffer through the capture membrane in reverse direction.

### Downstream PCR Detection

The eluent obtained from the filtration process underwent centrifugation at 7500 rpm for 10 minutes to yield a 20 µL purified and enriched bacteria sample, prepared for subsequent analysis. To account for detection of gram-positive bacteria, the final enriched bacteria samples underwent enzymatic lysis through treatment with a final concentration of 8U/mL mutanolysin at 37°C for 5 minutes. Subsequently, 10 µL of the resulting lysate was employed for PCR amplification. The PCR assay, conducted in a 25 µL reaction volume, comprised the following components: 1x SsoAdvanced Universal SYBR Green Supermix (Bio-Rad), 1 mg/mL bovine serum albumin (Thermo Scientific), 0.5x Evagreen (Bio-Rad), and 0.3 µM primer pairs targeting the nuc regions for S. aureus specificity, and tuf regions for E. coli specificity (refer to Table 1 for primer details). All primers were procured from Integrated DNA Technologies.

**Table 1:**
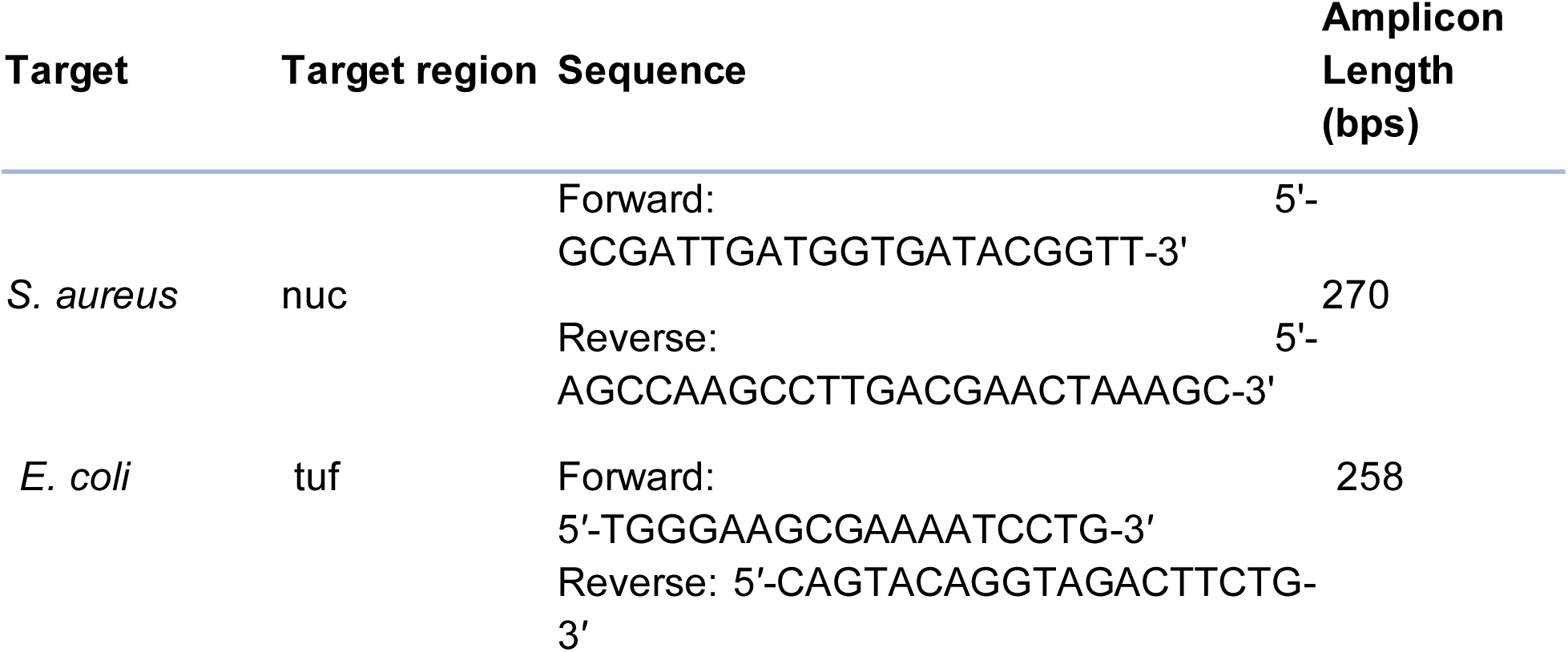
*S.aureus* and *E.coli* Specific Primer Design.

## Results and Discussion

### Overview of Workflow

Our blood processing protocol relies only on membrane filtration and can be entirely performed using syringes in 4 steps, thus obviating other specialized equipment. In our protocol, the blood sample containing bacteria was first 10 -fold diluted with water to induce osmolysis of red blood cells(RBCs) and peripheral blood mononuclear cells (PBMCs). The diluted blood was then filtered through a gradient filter (ranging from 35 μm to 2.5 μm) to reduce blood cells, large debris, and aggregates [22]. Next, the filtrate was incubated in CHAPs for 4 minutes and trypsin for 1 minute at room temperature to lyse residual RBCs. Subsequently, the sample was filtered through a 0.4 μm filter to capture the bacteria and separate them from the filtrate before a wash with water to further clean the sample. Finally, the captured bacteria could be recovered from the 0.4 μm filter by reversing the filter and performing an elution step with 0.1% tween 20 in 1 mL of water, readying them for downstream NAAT. Overall assay time is approximately 30 min (Fig. 1,Fig. S1).

**Figure 1.**
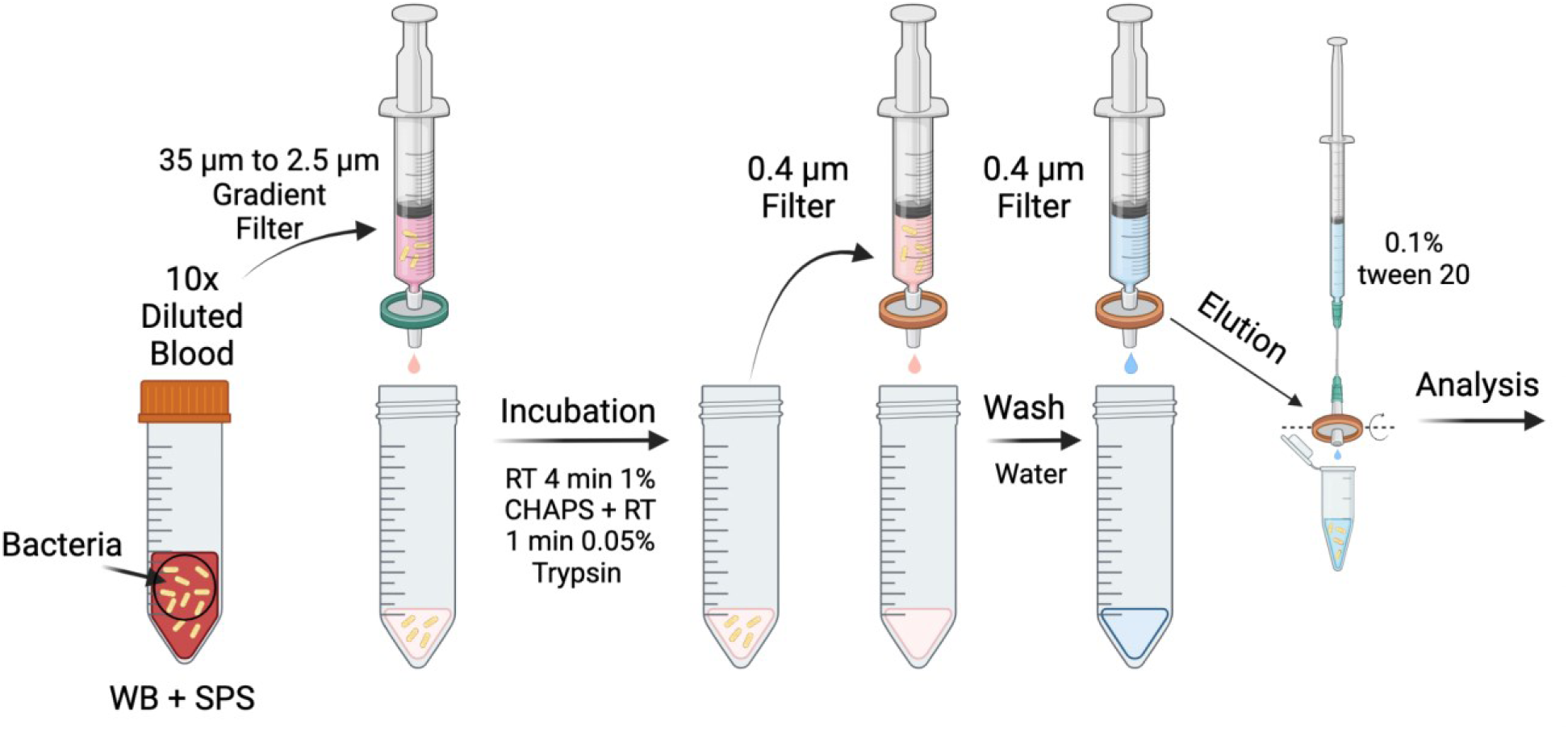
Overview of Filtration Assay. Human blood was pretreated with SPS, followed by bacteria inoculation and a 10-fold dilution in water to induce osmolysis. The diluted sample underwent filtration through a gradient filter (ranging from 35 μm to 2.5 μm). The resulting filtrate was incubated with CHAPs and trypsin to induce residual RBC lysis. Afterward, the sample was filtered through a 0.4 μm filter to capture bacteria, followed by a water flow-through for washing. The captured bacteria were eluted with 0.1% tween 20 in water for downstream analysis. The entire assay took approximately 30 minutes, and all analyses were conducted using qPCR reactions.

### Evaluation of Assay Conditions

In our pursuit of effective membrane filtration, we systematically examined different blood treatment conditions to ensure smooth passage through our dual-membrane system. Three distinct approaches were tested to assess their impact on filter performance. Initially, undiluted blood was found to easily clog the separation filter. Subsequent attempts involved a 10-fold dilution with DI water, inducing osmotic lysis of blood cells. While this addressed the separation filter issue, it unfortunately led to the capture filter’s obstruction. To overcome this challenge, we conducted a brief incubation of the filtrate from the first separation membrane with a combination of CHAPS and Trypsin at room temperature. These reagents played a crucial role in solubilizing proteins and lysing residual blood cells, enabling the unimpeded passage of the entire treated sample through the second membrane filter (Fig. 2A).

**Figure 2.**
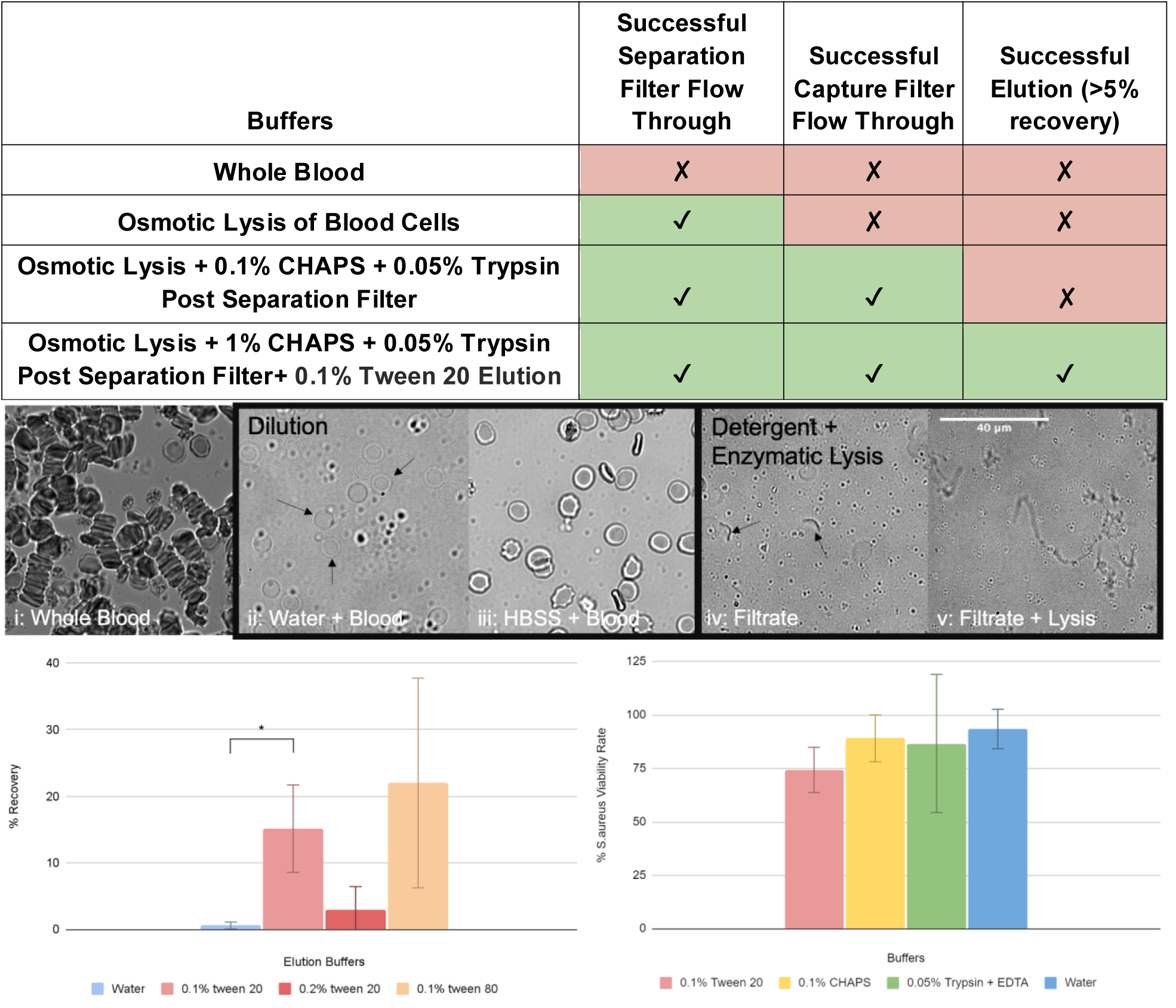
Development of Filtration Methods. **(A) Filtration Performance Analysis.** The filtration performance analysis comprises distinct stages. The filtration capability of whole blood is being evaluated through a 35 to 2.5 gradient filter for the separation of residual peripheral blood mononuclear cells (PBMC) from bacteria following a 10-fold dilution. Notably, the filtrate encounters difficulty flowing through the 0.4 μm filter for bacteria capture, a challenge overcome by introducing 0.1% CHAPS + 0.05% trypsin EDTA. The efficiency of bacterial elution from the 0.4 μm filter significantly improves upon the introduction of 0.1% tween 20. **(B) Blood Cell Lysis Assessment.** The osmotically lysed cells are observed under a brightfield microscope (40x objective) to evaluate the efficacy of both osmotic lysis and CHAPS+trypsin-based lysis. Noticeable distinctions are evident in both lysis conditions when compared to the unlysed sample. i.) SPS treated Whole Blood ii.) SPS treated whole blood diluted in water (osmotic lysis). Residual RBCs are indicated by arrows. iii.) SPS treated whole blood diluted in HBSS. iv.) Osmo-lysed blood filtered through a separation filter. Residual RBCs are indicated by arrows. v.) CHAPS and Trypsin treatment of osmo-lysed blood filtrate. All images undergo processing in ImageJ for visualization. **(C) Recovery Rate Assessment with Surfactant Additions.** In an attempt to enhance the recovery rate of the flow-through method, an assessment is conducted for 500 CFU S. aureus, captured on the 0.4 μm filter, using various surfactant additions (DEPC water, 0.1% Tween 20, 0.2% Tween 20, 0.1% Tween 80). Subsequent plating on 5% sheep blood TSA facilitates assessment. The recovery rates are determined as 0.6 ± 0.5%, 15.2 ± 6.6%, 2.9 ± 3.5%, and 22.0 ± 15.7%, respectively. Notably, 0.1% Tween 20 exhibits the highest statistically significant recovery rate in comparison to pure water (mean ± 1SD; n = 3; * p < 0.05). **(D) S. aureus Viability Testing in Each Buffer.** The viability of S. aureus after a 10-minute exposure to all buffers used in the assay (DEPC water, 0.1% Tween 20, 0.1% CHAPS, 0.05% Trypsin EDTA) is being assessed. The present viability is calculated by plating the buffer on TSA agar. The plating results are compared with the original CFU spike plating results. The viability assessments for 0.1% Tween 20, 0.1% CHAPS, 0.05% Trypsin EDTA, and DEPC water are calculated as 74 ± 10.6%, 89.1 ± 10.9%, 86.7 ± 32.2%, and 93.5 ± 9.2%, respectively. No significant difference is observed between all conditions (mean ± 1SD; n = 2; p > 0.05). All error bars visualized represent one standard deviation.

We employed microscopy to validate our observations at each stage of blood treatment. Without osmolysis, microscopic examination revealed stacked red blood cells (RBCs) in the blood sample (Fig. 2Bi), explaining the tendency to clog the separation filter. Osmolysis induced a noticeable reduction in RBCs, indicating partial lysis of blood cells (Fig. 2Bii). To further confirm osmolysis, we compared this with HBSS-diluted blood samples, where more RBCs were observed, affirming the occurrence of osmolysis (Fig. 2Bii and 2Biii). The efficacy of CHAPS+Trypsin-induced RBC lysis was assessed by imaging the separation filtrate before and after lysis buffer addition (Fig. 2Biv and 2Bv). Clear evidence of RBC lysis was observed throughout the treated solution. Based on these findings, our optimized blood treatment protocol involves a preliminary 10 -fold DI water dilution before the first separation filtration step and CHAPS and Trypsin treatment prior to the second capture filtration steps.

Utilizing S. aureus as our model bacterium, we trapped 100 CFU in the second membrane and assessed various elution conditions through plating. S. aureus was chosen for its significance as a Gram-positive bacterium causing both community-acquired and hospital-acquired bloodstream infections (BSIs), with mortality rates ranging from 10% to 30%, and higher rates for methicillin-resistant Staphylococcus aureus (MRSA) infections [23]. For elution, we tested Tween 20 at a concentration of 0.1% and 0.2%, while Tween 80 was tested at 0.1%. Results (Fig. 2C) indicated that 0.1% Tween 20 demonstrated statistical significance in increased CFU recovery, achieving rates of 15-20%. We acknowledge that vortexing the second filter membrane could enhance recovery (Fig. S5). However, prioritizing a straightforward protocol, we opted for direct elution without the vortexing step.

We also assessed the integrity of bacteria when exposed to various buffers in our protocol. S. aureus and E. coli were subjected to CHAPS, Trypsin, Tween 20, and DI water (used as the wash buffer) for 10 minutes, followed by plating to evaluate their integrity. Results (Fig. 2C) demonstrated the recovery of at least ∼75% of S. aureus cells after exposure, indicating their overall integrity. Similarly, E. coli, a Gram-negative bacterium responsible for approximately 5.6% of bacteroides species-related bloodstream infections [24], displayed comparable results. The mortality rate associated with E. coli BSIs varies (16.9% to 33.9%) based on factors such as antimicrobial resistance and the patient’s health status [24]. Our protocol maintained the integrity of the majority of E. coli cells (Fig. S4).

### Sensitive Detection of Two Causative Bacteria

We coupled our blood processing protocol with downstream PCR for sensitive detection, focusing on S. aureus and E. coli as model organisms due to their prevalence in bloodstream infections (BSI) [25]. Both species were spiked at approximately 100, 50, and 20 CFU into blood samples. To ensure precision, we plated the bacteria suspension before spiking. Benchtop PCR was performed using S. aureus-specific PCR primers from Brakstad et al. [26] and E. coli-specific PCR primers from Maheux et al. [27]. For S. aureus, clear PCR curves were observed in all spiked samples, with non-amplified fluorescence signals in negative controls. The PCR Cq values were 34.73 ± 1.69, 36.23 ± 0.61, 39.46 ± 3.42, and 60 (no amplification) for 92 ± 23 CFU, 42 ± 6 CFU, 12 ± 5 CFU, and negative controls, respectively (Fig. 3A), indicating successful detection of S. aureus at sepsis-relevant bacterial loads. Similarly, for E. coli, clear PCR curves were observed in all spiked samples and some negative controls. The PCR Cq values were 34.82 ± 0.79, 36.30 ± 1.87, 37.85 ± 1.52, and 53.61 ± 9.03 (background signal) for 55 ± 18 CFU, 28 ± 9 CFU, 11 ± 3 CFU, and negative controls, respectively (Fig. 3B; n = 3). Despite undesired amplification in negative controls, the Cq values remained significantly distinct from those of the samples (p < 0.05), supporting the successful detection of E. coli at sepsis-relevant bacterial loads.

**Figure 3:**
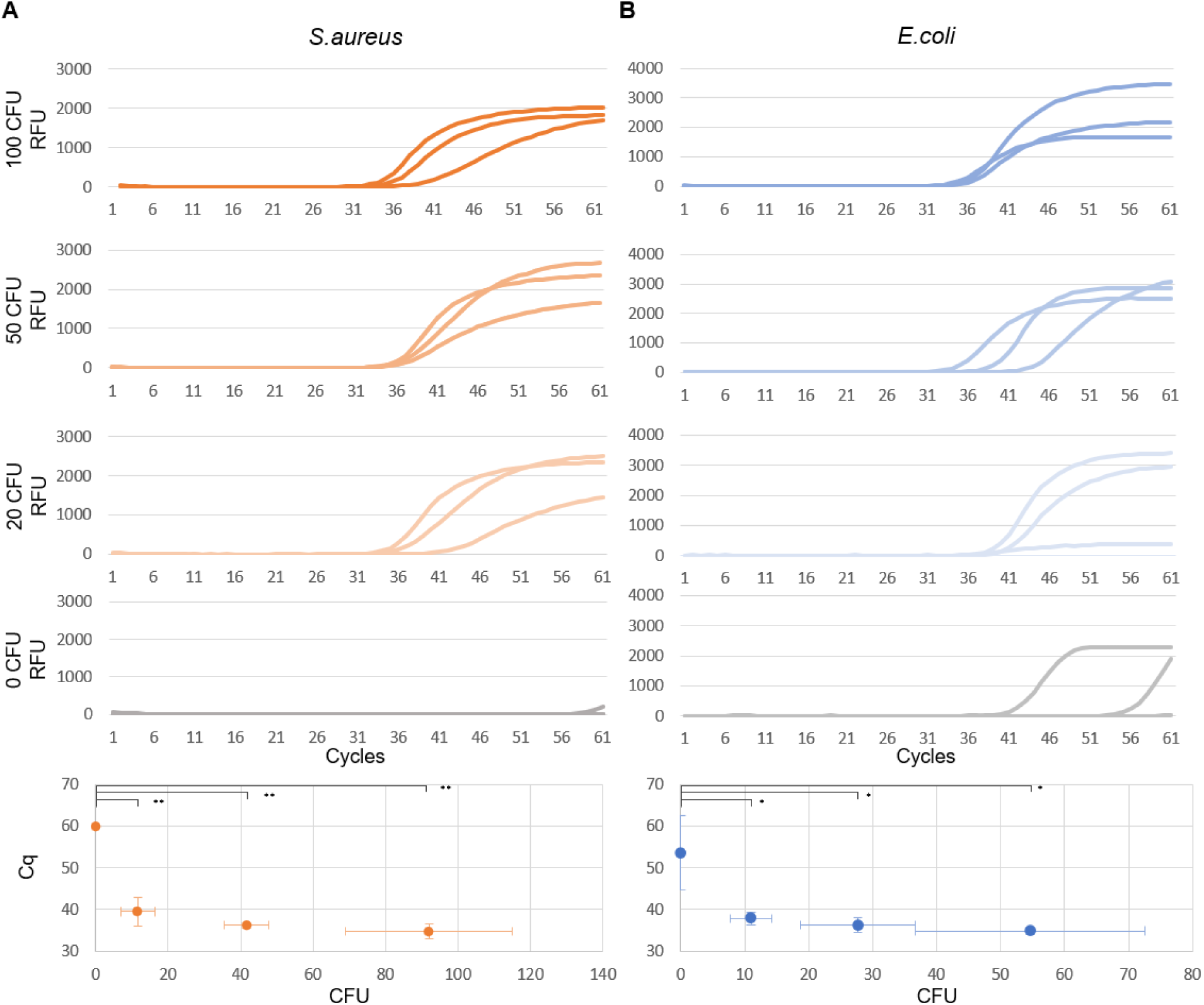
qPCR Analysis of Filtration Sample. This figure illustrates the results of qPCR analysis for E. coli and S. aureus, obtained through a series of steps involving blood treatment with SPS, bacterial spiking, and subsequent filtration, as previously outlined. Plating the bacterial suspensions before spiking into blood ensures more precise colony counts. **(A) S. aureus Detection.** For S. aureus, we analyze samples at approximate concentrations of 100 CFU, 50 CFU, 20 CFU, and 0 CFU. Replicated quantification cycle (Cq) values are 34.73 ± 1.69, 36.23 ± 0.61, 39.46 ± 3.42, and 60 (i.e., no amplification at cycle 60) respectively (mean ± 1SD; n = 3; ** p < 0.01). Clear detection of S. aureus is evident with statistical significance. **(B) E. coli Detection.** Similarly, we perform E. coli detection for approximate concentrations of 100 CFU, 50 CFU, 20 CFU, and 0 CFU. The resulting average Cq values are recorded as 34.82 ± 0.79, 36.30 ± 1.87, 37.85 ± 1.52, and 53.61 ± 9.03 respectively (mean ± 1SD; n = 3; * p < 0.05). Clear detection of E. coli is observed with statistical significance.

## Discussion

In this study, we present a new blood processing method that allows for the culture-free, rapid, and low-volume isolation of pathogenic bacteria from whole blood. Our method combines osmolysis, separation membrane filtration, detergent and enzymatic lysis, and bacteria isolation, which can be executed using minimal specialized equipment. One of the major achievements of our work is the elimination of the need for pre-culture or post-culture steps. This culture-free approach not only accelerates the diagnosis process but also minimizes the risk of altering bacterial populations during culturing, which can impact clinical decisions. Another strength of our method lies in its requirement for small blood sample volume (currently 0.5 mL or below), which can potentially facilitate use with finger-prick blood samples. This feature not only simplifies the sampling process but also broadens the applicability of our assay, making it suitable for a wider range of patients, including pediatrics, where obtaining large blood volumes can be challenging. Moreover, our approach directly addresses the limitations of traditional blood culturing methods, which typically require up to 20 mL of blood to provide an accurate detection of bloodstream infections [28].

We envision several areas for improvement. First, in this study, we focused on two causative bacteria, S. aureus and E. coli, due to their prevalence in bloodstream infections. We selected S. aureus as a model bacterium due to its challenging characteristics in blood detection. Its gram-positive nature makes it resistant to thermolysis, posing difficulties for PCR [29]. Additionally, its attachment to red blood cells introduces further challenges, especially when employing methods such as RBC sedimentation [11,30]. Moreover, the strong binding to IgG, facilitated by the surface protein A present in S. aureus, adds another layer of complexity to the detection process [31,32]. However, we recognize the importance of testing a broader range of bacteria to assess the general utility of our protocol. We also envision the possibility of extending this method to test fungal pathogens, further broadening its clinical applications. Second, while we consistently achieved the detection of 20 CFU and even down to approximately 10 CFU of bacteria using benchtop PCR, the quest for single-copy sensitivity remains a challenge. To improve sensitivity, a feasible strategy would be to expand the blood sample volume. Our focus on small blood volumes for proof-of-concept serves specific scenarios, such as pediatric applications, but enlarging the membrane filter areas could allow for more extensive sample sizes. To further enhance the recovery from the capture membrane, we have explored alternative elution methods, such as filter removal followed by vortexing and surfactant elution, which have shown promise in our preliminary testing with additional 6% recovery (Fig S5). In the future, further optimization in elution methods in tandem to a specialized filter membrane holder design may be considered to overcome this challenge. Third, we combined our blood processing workflow with benchtop PCR as an initial demonstration. We envision appending other detection methods such as rapid, magnetofluidic-enabled PCR [33–35] to achieve fast turnaround, or digital PCR-HRM [36] to achieve polymicrobial detection. Finally, plans for antimicrobial susceptibility testing (AST) represent a crucial step toward comprehensive clinical utility. Future research should focus on refining the elution step, exploring alternative filter materials and pore architectures, and expanding the method’s applicability to a broader range of pathogens.

In conclusion, our research introduces a versatile and innovative method for the isolation of pathogenic bacteria from whole blood. The implications of our findings are profound, offering a rapid and culture-free approach that simplifies the diagnostic process for bloodstream infections. With the potential for direct integration into clinical workflows, especially when coupled with magnetofluidic PCR for point-of-care testing in resource-limited environments, our method holds promise in advancing clinical microbiology. Although challenges remain, such as achieving single-copy sensitivity, expanding blood sample volumes, and further testing with various pathogens, our work represents a significant step toward a more efficient and accessible approach to diagno sing life-threatening bacterial infections. The future of sepsis diagnosis may indeed be simplified, rapid, and accessible to a wider range of patients, thanks to innovative techniques such as the one presented in this study.

## Data Availability

All data produced in the present study are available upon reasonable request to the authors.

## Supplementary Data

**S1:**
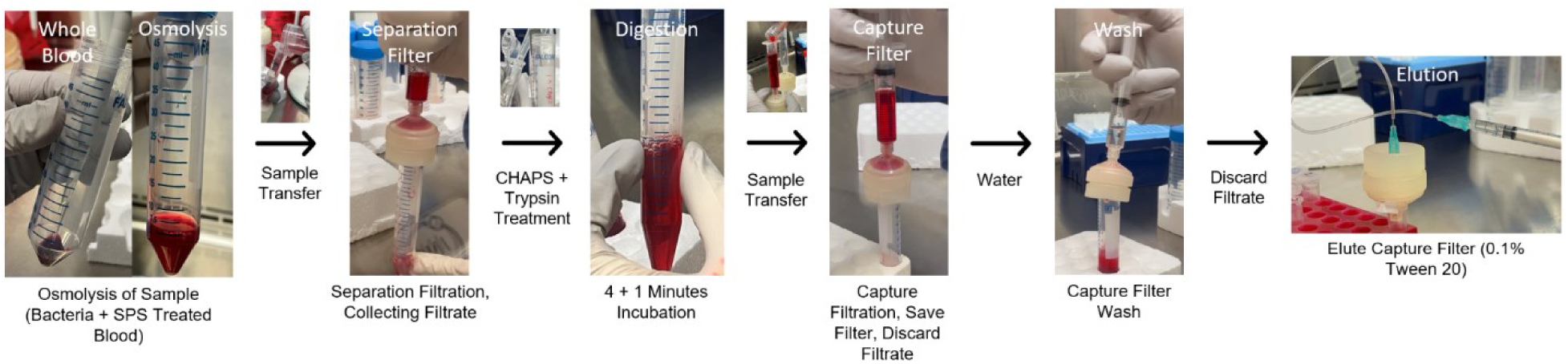
Filtration Workflow. This figure depicts the filtration workflow, providing a step-by-step guide. The blood sample, treated with SPS and water, is inoculated with bacteria, initiating the subsequent filtration protocol as previously outlined.

**S2:**
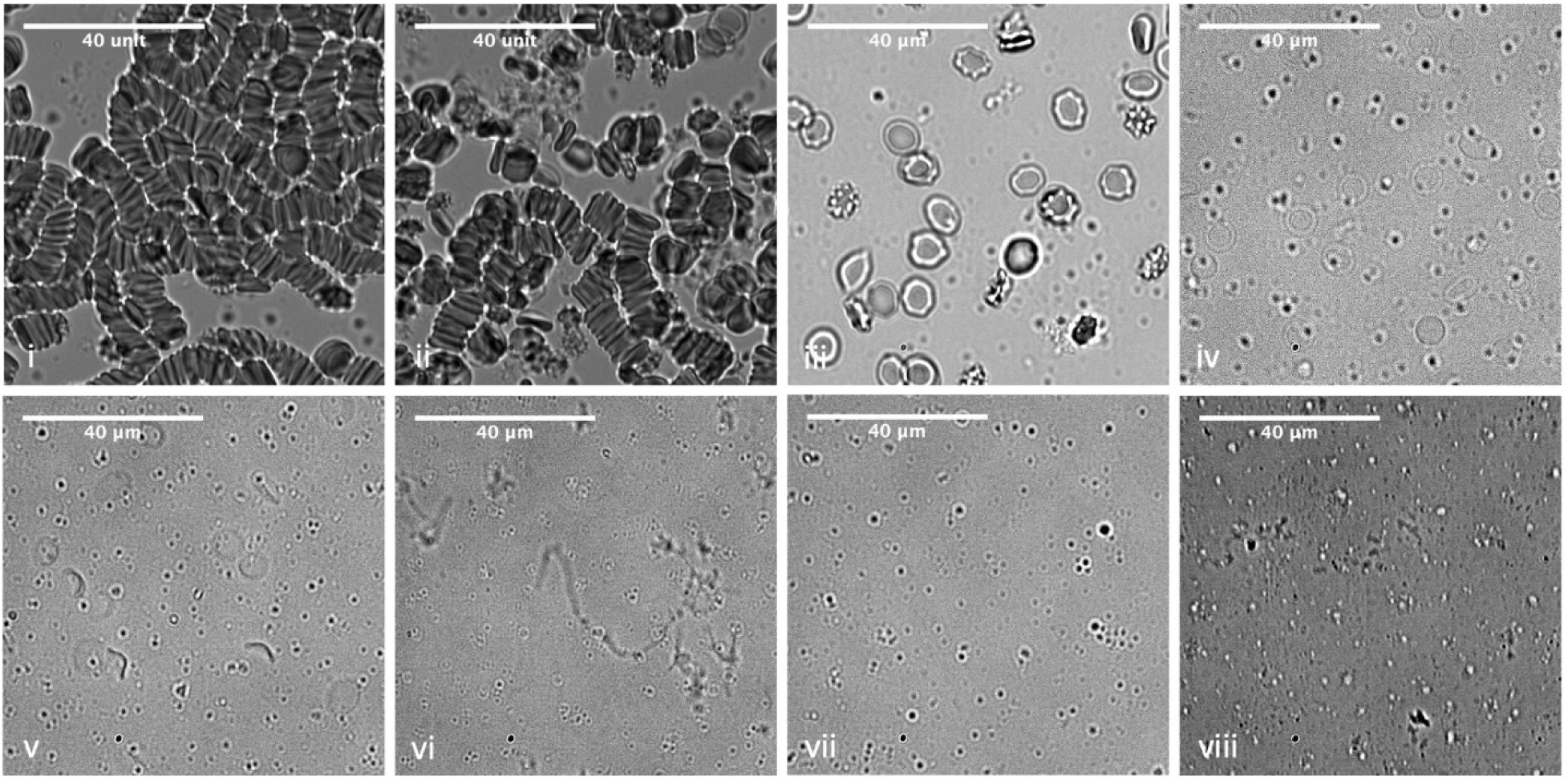
Assessment of blood under each step of filtration methods. The blood sample was observed under a brightfield microscope (40x objective) to assess the lysis capability of osmotic lysis and CHAPS+Trypsin-based lysis. i.) Whole Blood ii.) SPS treated whole blood iii.) SPS treated whole blood diluted in HBSS. iv.) SPS treated whole blood diluted in water (osmotic lysis). v.) Osmo-lysed blood filtered through a separation filter. v.) CHAPS and Trypsin treatment of osmo-lysed blood filtrate. vi) Capture filter eluent. vii) Capture filter filtrate. All images were processed in ImageJ for visualization.

**S3.**
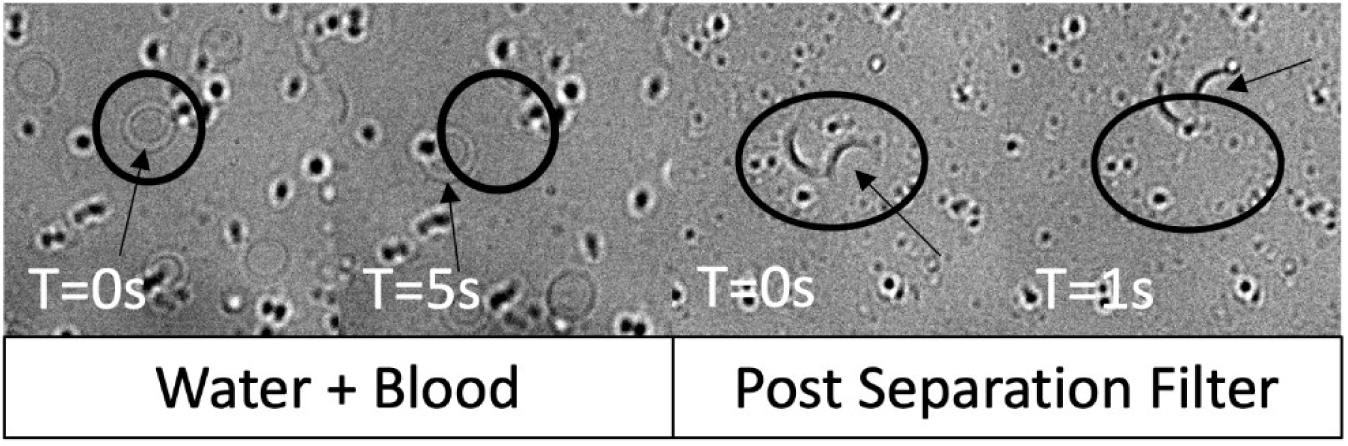
Identification of RBC from the Background. To discern residual blood cells from the background, a sequence of images was captured at 0.1-second intervals, totaling 50 images at a 40x objective. As indicated by the arrow, the movement of red blood cells (RBCs) is observable, contrasting with the stationary black background dots.

**S4.**
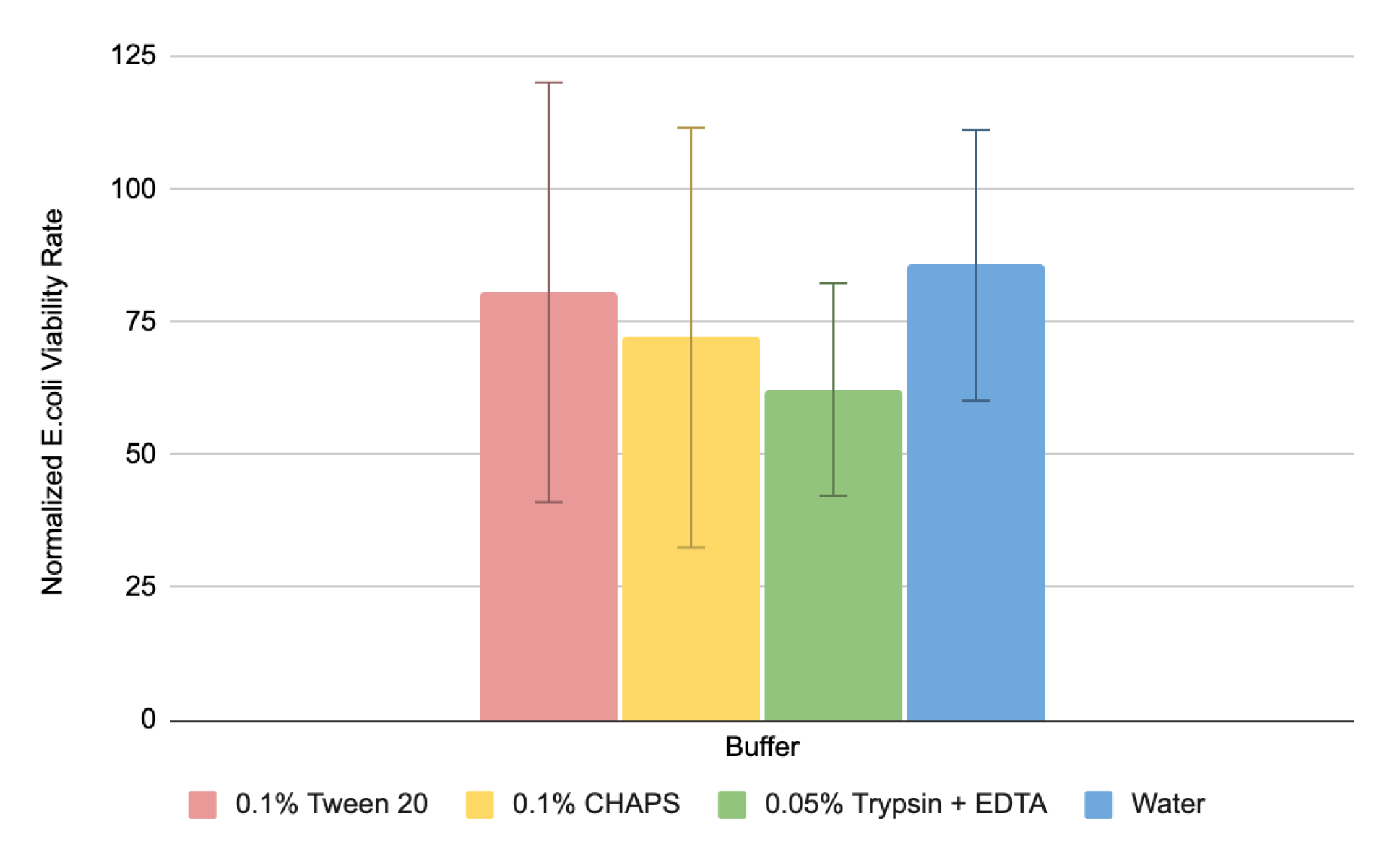
E. coli Viability Testing in Each Buffer. We assess the viability of E. coli after a 10-minute exposure to all buffers used in the assay (DEPC water, 0.1% Tween 20, 0.1% CHAPS, 0.05% Trypsin EDTA). The present viability was calculated by plating the buffer on TSA agar, and the results were compared with the original CFU spike plating results. The viability assessments for 0.1% Tween 20, 0.1% CHAPS, 0.05% Trypsin EDTA, and DEPC water are calculated as 80.6 ± 43.2%, 72.1 ± 39.5%, 62.3 ± 20.0%, and 85.7 ± 25.5%, respectively. No significant difference is observed between all conditions (n = 2).

**S5.**
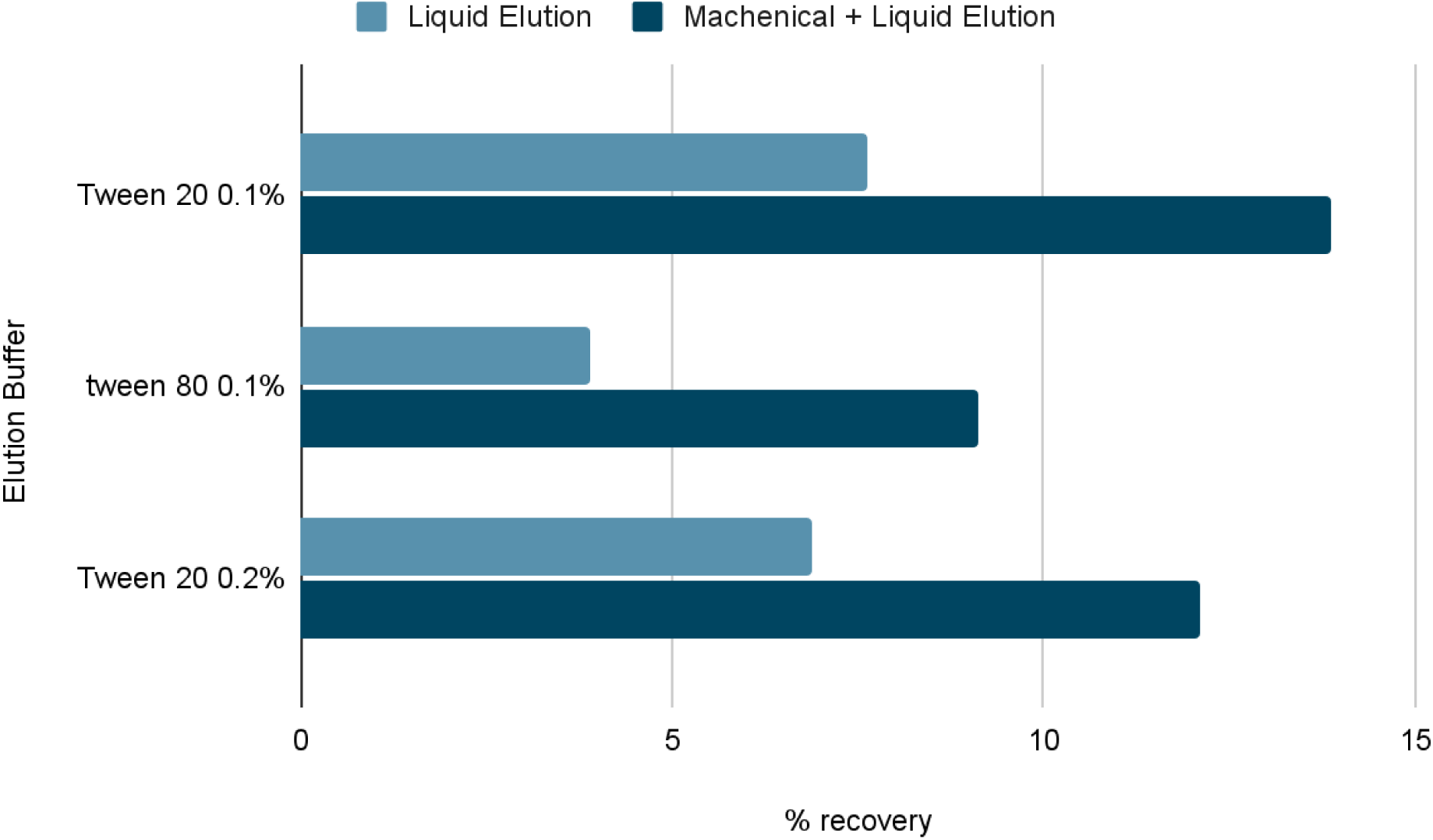
Exploring Alternate Bacteria Elution Methods. In addition to the previously proposed flow-through method, we investigate an alternative mechanical elution method by removing the capture filter paper and vortexing it in the elution buffer. This results in an increased recovery of 6.3%, 5.2%, and 5.2% for Tween 20 0.1%, Tween 80 0.1%, and Tween 20 0.2% elution buffer, respectively.

## Notes

### Competing Interest Statement

The authors have declared no competing interest.

### Funding Statement

This study was funded by NIH grant (R01AI137272).

